# Evaluating the design and implementation fidelity of an adapted Plan-Do-Study-Act approach to improve health system performance in a Nigerian state

**DOI:** 10.1101/2020.08.31.20185322

**Authors:** Ejemai Amaize Eboreime, John Olajide Olawepo, Aduragbemi Banke-Thomas, Rohit Ramaswamy

**Affiliations:** Department of Medicine, Faculty of Medicine and Dentistry, University of Alberta, Edmonton, AB, Canada; Department of Planning, Research and Statistics, National Primary Healthcare Development Agency, Abuja, Nigeria; Department of Environmental and Occupational Health, School of Public Health, University of Nevada Las Vegas, USA; Department of Health Sciences, Bouve College of Health Sciences, Northeastern University, Boston, USA; Department of Health Policy, London School of Economics and Political Science, London, UK; Gillings School of Global Public Health, University of North Carolina at Chapel Hill, Chapel Hill NC, USA

**Keywords:** Quality Improvement, Implementation Science, PDSA, Health System, Nigeria, program design

## Abstract

**Background:** The Plan-Do-Study-Act (PDSA) cycle is fundamental to many quality improvement (QI) models. For the approach to be effective in the real-world, variants must align with standard elements of the PDSA. This study evaluates the alignment between theory, design and implementation fidelity of a PDSA variant adapted for Nigeria’s health system performance improvement.

**Methods:** An iterative consensus building approach was used to develop a scorecard evaluating new conceptual indices of design and implementation fidelity of QI interventions (design and implementation index, defects and gaps) based on Taylor’s theoretical framework.

**Results:** Design (adaptation) scores were optimal across all standard features indicating that design was well adapted to the typical PDSA. Conversely, implementation fidelity scores were only optimal with two standard features: prediction-based test of change and the use of data over time. The other features, use of multiple iterative cycles and documentation had implementation gaps of 17% and 50% respectively.

**Conclusion:** This study demonstrates how both adaptation and implementation fidelity are important for success of QI interventions. It also presents an approach for evaluating other QI models using Taylor’s PDSA assessment framework as a guide, which might serve to strengthen the theory behind future QI models and provide guidance on their appropriate use.

## Introduction

The Plan-Do-Study-Act (PDSA) cycle (and its concept of iterative tests of change) is fundamental to both the fields of Quality Improvement (QI) and Implementation Science. Whereas Implementation Science is primarily concerned with promoting the “integration of evidence-based interventions into health care policy and practice”, QI science concentrates on “systematically and rigorously exploring ‘what works’ to improve quality in healthcare” (Health Foundation, 2011). Both disciplines place an emphasis on improving care using a systems-level approach. Both also overlap by considering concepts such as context, the organizational and system actors (providers, staff, patients, and administrators), processes and outcomes (Williams, 2019).

Many popular QI models such as the Model for Improvement, Lean and Six Sigma (Schroeder *et al.*, 2008; Norman *et al.*, 2013; Taylor *et al.*, 2013; Reed and Card, 2016) incorporate some version of PDSA in their methodologies to test improvement solutions on a small scale before scaling up. In the field of Implementation Science, PDSA is similarly used to adapt implementation strategies to local contexts (Fixsen *et al.*, 2005; Powell *et al.*, 2015). Whereas Implementation Science considers the PDSA as a method or strategy, the approach is viewed as an intervention in the science of improvement (Koczwara *et al.*, 2018). A recently proposed model integrating QI and implementation science, called the Model for Improvement and Implementation (Ramaswamy, Johnson and Hirschhorn, 2018) explicitly situates PDSA as a process at the intersection of both fields, necessary for both context specific intervention development (QI) and adaptation of IS strategies.

Moreover, PDSA cycles align well with models for decision-making in complex systems with non-linear processes that are unpredictable and that display emergent behaviour (WHO, 2009). In such systems, a “probe, sense, respond” approach is recommended, where decision makers initiate small experiments to test how the system behaves before engaging in large scale analysis and intervention (Snowden and Boone, 2007). As organizations such the World Health Organization and the World Bank recommend QI to address larger scale problems such as health systems strengthening to improve global quality of care (Kieny *et al.*, 2018), the use of PDSA is likely to continue to expand.

The use of PDSA to find local solutions to solve complex improvement and implementation problems offers great promise (Muczyk, 2004; Sales *et al.*, 2006), and this approach can be considered to be a best practice in developing quality improvement interventions. However, even though PDSA provides a rigorous experimental approach to improvement, it has not been always been used effectively to bring about outcomes (Reed and Card, 2016). The reason for this is there is no standard protocol or formula for using PDSAs. This in itself is not a shortcoming – by definition, since PDSA cycles are intended to drive learning based on generating hypotheses about how to change a system, a PDSA approach should be deeply embedded in the systems in which implementation and improvement work takes place and its particular application should depend on the context of the system as several researchers have pointed out (Øvretveit, 2011; Leis and Shojania, 2017; Ramaswamy *et al.*, 2018). However, this assumes that those engaged in improvement or implementation adhere to the primary principles of the PDSA such as developing change hypothesis, designing multiple tests of increasing complexity, collecting and analysing data and making appropriate updates to the change hypothesis. As demonstrated by Taylor and colleagues (Taylor *et al.*, 2013) who applied an evaluation framework to assess gaps in the use of PDSA cycles to improve health care, this is not often the case. Despite the theoretical promise of PDSA, it often fails in execution.

That being said, the PDSA approach still remains the best theoretical model for designing quality improvement interventions, and therefore it is appropriate to test other models designed to improve quality, against the PDSA approach as a standard. While any quality improvement approach needs to be implemented well, there is an additional barrier to success if the theory underlying the approach is untested or flawed. At the same time, as mentioned above, PDSA is not an inviolable formula, and therefore needs to be adapted for use in various settings. In this paper, we evaluate a quality improvement model called Diagnose-Intervene-Verify-Adjust (DIVA), adopted in Nigeria and other low and middle-income countries (LMICs) to improve planning processes in health systems.

DIVA was developed, introduced, tested and implemented in several LMICs by United Nations Children’s Fund (UNICEF) (UNICEF, 2012). The model seeks to improve processes such as routine planning cycles and the use of data by local stakeholders and policy/decision makers as suggested by the Lancet Global Health Commission on High Quality Health Systems (HQSS) (UNICEF, 2012; O’Connell and Sharkey, 2013). Typically, operational planning occurs annually. DIVA strengthens the annual planning cycles by introducing quarterly iterations for monitoring and improvement of the annual plan. Further, outputs from DIVA (such as proposed implementation strategies for health interventions) feed into subsequent operational planning cycles. DIVA also builds the capacity of district/local government health teams, communities and other local stakeholders to identify bottlenecks and barriers to the planning process. The model provides tools to address these barriers, take action and be accountable for service delivery within their jurisdiction towards improving effective coverage, quality and impact of health interventions (UNICEF, 2012; O’Connell and Sharkey, 2013).

Operationally, DIVA is a four-step model in which ‘Diagnose’ identifies bottlenecks to health interventions; ‘Intervene’ develops and implements action plans to resolve identified constraints; while ‘Verify/Adjust’ tracks the results of the action plans and recommends real-time data driven adjustments to the plan (Figure 1). DIVA uniquely doubles as a Quality Improvement method as well as an implementation strategy for evidence informed planning of interventions in complex health systems. The model was developed in response to the observation that district health planning in many LMICs is commonly fragmented across various actors and vertical programmes, and often not informed by contextual evidence. The theory of change behind the model is that evidence-informed bottleneck analysis and integrated intervention planning using DIVA, will improve efficiency of resource allocation and utilization as well as health system performance indices (UNICEF, 2012, 2016).

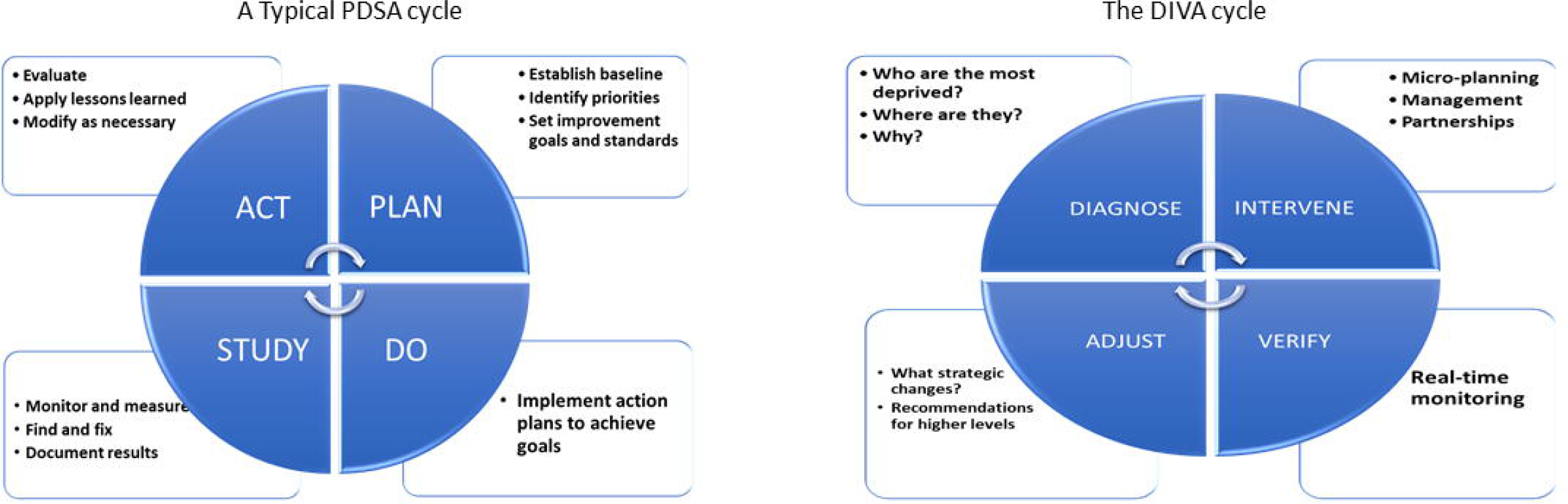

Research, published elsewhere (Eboreime, Nxumalo, *et al.*, 2019) showed that the DIVA model can potentially improve health system performance, but that effective implementation is critical. Other evaluations in Nigeria, Ghana and Uganda have also demonstrated that the tools and methods of DIVA are effective in evidence-informed planning of health interventions (O’Connell and Sharkey, 2013; Baker, Okuga, *et al.*, 2015; Baker, Peterson, *et al.*, 2015; Henriksson *et al.*, 2017; Eboreime, Nxumalo, *et al.*, 2018) but outcomes have exhibited significant variability (Eboreime, Eyles, *et al.*, 2018; Eboreime, Nxumalo, *et al.*, 2018). To provide guidance to countries seeking to enhance the effectiveness of DIVA, it would be useful to assess both the theoretical underpinning and the implementation fidelity of DIVA. This will help practitioners and policy/decision makers evaluate whether the failure of DIVA to achieve outcomes results from a failure in its design (i.e. the underlying premise of DIVA is flawed) or whether DIVA is a context-appropriate adaptation of the principles of PDSA, and failures in its performance can be attributed to poor implementation fidelity. If the latter is the case, strategies to improve the implementation of DIVA can be informed by the substantial body of knowledge that already exists about how to conduct PDSAs well. Specifically, in this paper, we seek to answer the following research questions:

1. To what extent can DIVA be considered to be an adaptation of PDSA?
2. To what extent does the implementation of DIVA in Kaduna state, Nigeria exhibit fidelity to its design?

## Materials and Methods

### Study context

Nigeria operates a decentralized three-tier health system governance structure. The federal level is responsible for overarching policy thrusts including programme design, the state ministries of health are largely responsible for subnational coordination and translation, while the local governments are mostly the implementers (FMOH, 2016; E. A. Eboreime *et al.*, 2017). DIVA was introduced nationwide in 2012, most states experienced implementation failure with only Kaduna sustaining implementation as at the time of this study (Eboreime, Eyles, *et al.*, 2018; Eboreime, Nxumalo, *et al.*, 2018).

### Theoretical Framework

To answer the first research question, we used the theoretical framework developed by Taylor and colleagues (3) mentioned earlier to assess the quality of implementation of PDSA cycles in 47 studies in healthcare settings. This framework uses five features to test the theory and the practice of the approach. These features are the use of Iterative cycles; prediction-based test of change, small-scale testing; use of data over time, and documentation of processes and outcomes for learning (Table 1). Given the use of the DIVA implementation for adjusting the annual plan on a quarterly basis, one of these characteristics (small scale testing) was not relevant for DIVA, because the steps of the DIVA process were applied to the same plan every quarter. We adapted this framework first to evaluate whether the design guidelines for DIVA include these key features. We used our findings from this evaluation to develop the assessment questions to evaluate the fidelity of implementation.

**Table 1:** Taylor’s theoretical framework based on key features of the plan–do–study–act (PDSA) cycle(Source: (Taylor *et al.*, 2013))

| Feature of PDSA | Description of feature |
| --- | --- |
| Iterative cycles | To achieve an iterative approach, multiple PDSA cycles must occur. Lessons learned from one cycle link and inform cycles that follow. Depending on the knowledge gained from a PDSA cycle, the following cycle may seek to modify, expand, adopt or abandon a change that was tested. |
| Prediction-based test of change | A prediction of the outcome of a change is developed in the 'plan' stage of a cycle. This change is then tested and examined by comparison of results with the prediction. |
| Small-scale testing | As certainty of success of a test of change is not guaranteed, PDSAs start small in scale and build in scale as confidence grows. This allows the change to be adapted according to feedback, minimises risk and facilitates rapid change and learning. |
| Use of data over time | Data over time increases understanding regarding the variation inherent in a complex healthcare system. Use of data over time is necessary to understand the impact of a change on the process or outcome of interest. |
| Documentation | Documentation is crucial to support local learning and transferability of learning to other settings. |

### Data collection

Data for the overall implementation research of which this work is a component were obtained from content analyses of 39 policy documents, 15 in-depth interviews and an embedded process observation between 2012 and 2016 to identify the design and implementation processes of DIVA in Nigeria. The documents were identified from the researchers’ knowledge (one author was an embedded researcher) and interviews of four national level subject matter experts who contributed to the initial design and implementation processes. The documents include policy guidelines, a DIVA implementation guide and implementation reports obtained at both the design phase (national) and implementation phases (subnational). This sub-study uses data related to the implementation of DIVA in Kaduna state because this is the only state still sustaining the implementation of DIVA since its design and deployment in 2012 (Eboreime, Eyles, *et al.*, 2018; Eboreime, Nxumalo, *et al.*, 2018).

### Analytic parameters

We developed the following indicators to perform our evaluation:

1. *Adaptation score/index:* This is a measure of the extent to which the steps of DIVA is consistent with the applicable elements of PDSA. It assesses the conceptual similarity between the standard improvement model and its local adaptation.
2. *Implementation score/index:* This is a measure of the extent to which the adapted model was implemented in the real world. The implementation index may be considered to be a measure of adherence to how DIVA was intended to be used, which is a dimension of implementation fidelity (James Bell Associates, 2009).

Figure 2 is an illustration of these concepts.

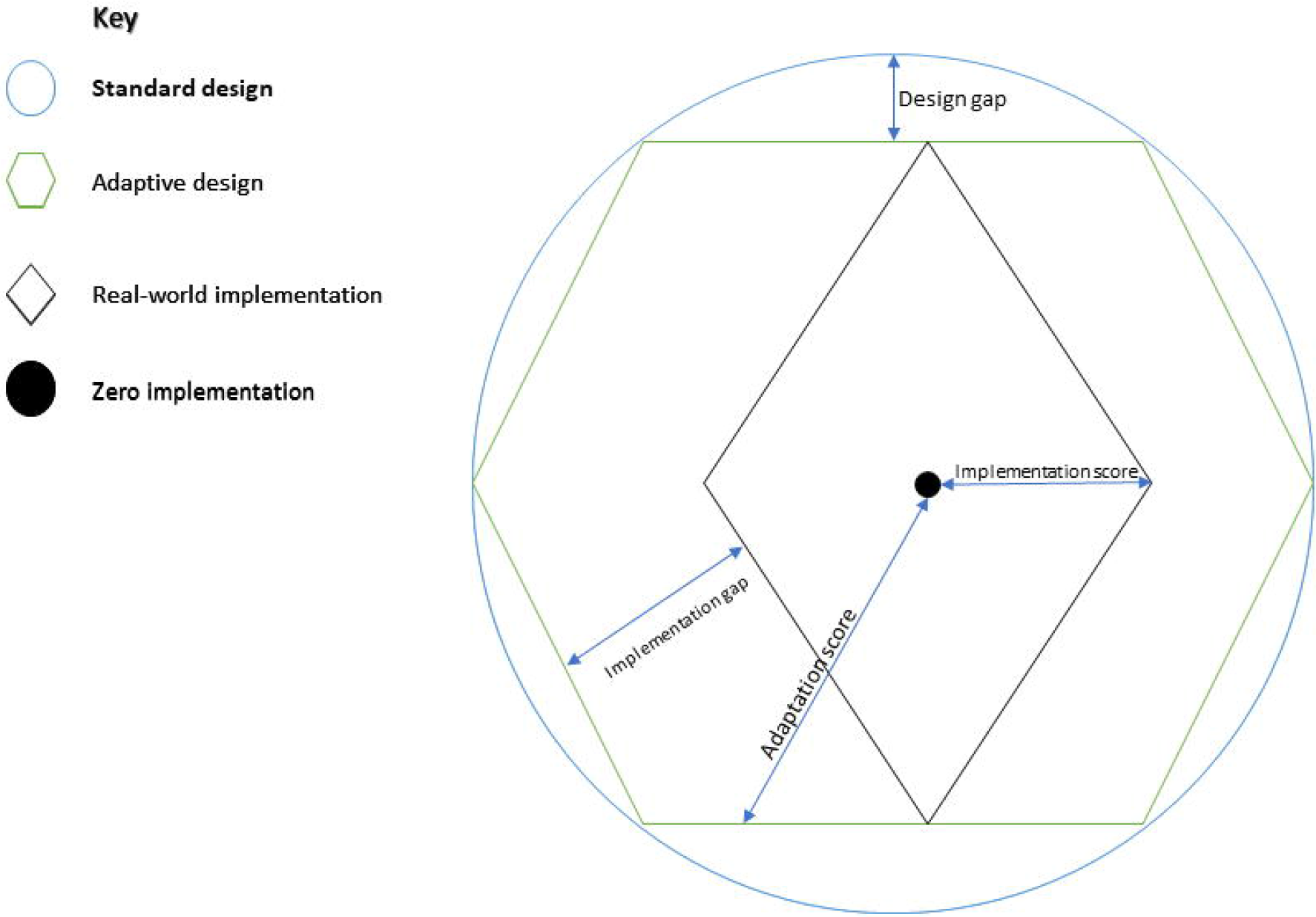

Taylor’s framework used 13 questions to assess the implementation fidelity of PDSAs, which are paraphrased in Table 2. To apply this framework to the design of DIVA, an adapted guide was created. The design evaluation helped to clarify the key features of the DIVA model and the extent to which these features were aligned with the key steps of a PDSA approach. Based on the results, an implementation fidelity evaluation questionnaire was developed. This questionnaire is shown in Table 3.

**Table 2:**
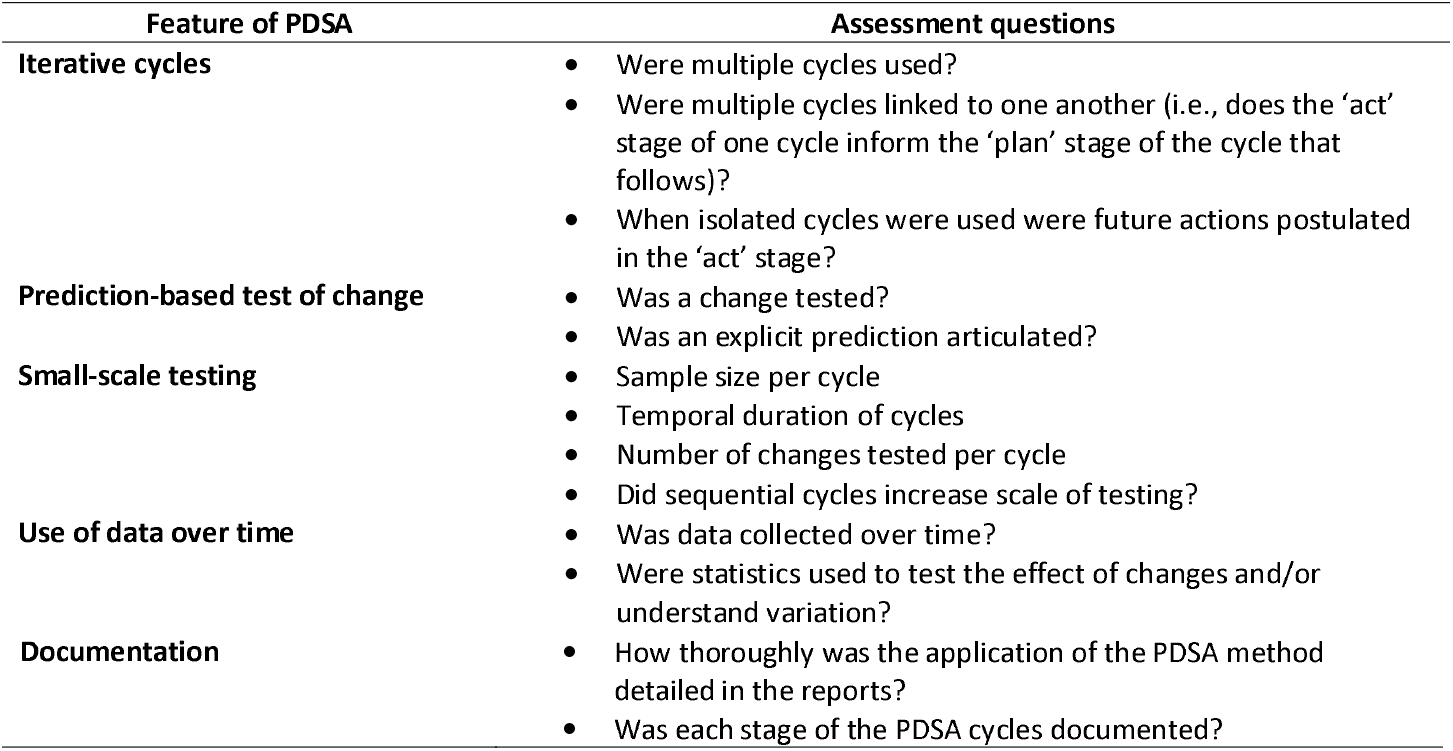
Taylor’s PDSA assessment questions

**Table 3:** Data analysis tool

| Feature of PDSA | Analysis criteria |  | Response scale for each question (Scores: 0/1/2) |
| --- | --- | --- | --- |
|  | Design evaluation | Implementation evaluation |  |
| Iterative cycles | Does the DIVA model incorporate the principle of multiple cycles?<br>Are multiple cycles linked to one another (i.e., does the 'adjust' stage of one cycle inform the 'diagnose' stage of the cycle that follows)? | Were multiple cycles used?<br>Were multiple cycles linked to one another (i.e., does the 'adjust' stage of one cycle inform the 'diagnose' stage of the cycle that follows)? | No/partly/yes |
| Prediction-based test of change | Does DIVA depend on well-defined theory of change?<br>Does the use of the model require explicit predictions that are tested in the "intervene" step? | Was a theory of change articulated prior to the implementation?<br>Was an explicit prediction made in each step? | No/partly/yes |
| Small-scale testing | Sample size per cycle<br>Temporal duration of cycles<br>Number of changes tested per cycle | Not applicable | Decrease/ same/ increase |
| Use of data over time | Does the DIVA model require the collection and display of data over time? | Was data collected over time? | No/partly/yes |
| Documentation | Does the design require detailed reporting of the application of the DIVA model in each quarter?<br>Does the design require detailed reporting of each step of the DIVA process in each quarter? | Was the application of the DIVA method detailed in the reports?<br>Was each stage of the DIVA cycle(s) documented? | No/partly/yes |

### Data analysis

To assess the level of alignment between the data and standard features, we created a scorecard. The scorecard assigned quantitative scores that measured the extent to which the reported or documented design or implementation activity aligned with the theoretical framework. An iterative consensus building approach was used to discuss and finalize scores. Fowles’ Delphi process guided the analysis(Van Wyk, 1980). This process mirrors the approach used in evaluating the deployment and installation processes of DIVA in the other studies (Eboreime, Eyles, *et al.*, 2018; Eboreime, Nxumalo, *et al.*, 2018).

A team of three PDSA/ DIVA subject matter experts was constituted and a process facilitator was appointed. The first round of questionnaires was developed using the questions from Taylor’s framework. These questions were tested and adapted to the study context.

Each question in the design and implementation was evaluated on a 3-point ordinal scale (No = 0, Partly = 1, Yes = 2). ‘No’ and ‘Yes’ were attributed to absolute non-compliance and absolute compliance respectively, whereas a ‘partly’ was assigned to partial compliance. The three evaluators with expertise and knowledge of the PDSA and DIVA independently reviewed the qualitative data and assigned a score to each question. Inter-rater variances were resolved using a consensus building discussions. The rules were that disparities between each rater’s scores on any question must not exceed one point. Thus, any rating disparity greater than one point was discussed and re-evaluated in the next round. The underlying assumption was that a one-point difference may represent random variation which could be harmonized by calculating a mean. However, a two-point disparity may be indicative of a significant interpreter difference which may need more clarification. The mean was used as the final score for each question. Each question carried an equal weight per standard feature. The mean score for each feature was computed and composite scores for each feature were created by dividing by a factor of 2 expressed in percentage values ranging from 0% (Zero implementation) to 100% (full implementation/full design alignment).

## Results

Table 4 presents the scores for design and implementation fidelity of DIVA on each evaluation criterion. Our design evaluation indicated that all the applicable characteristics of PDSA were part of the DIVA design, and therefore DIVA can be considered an adaptation of the PDSA model. We could therefore use the same questions we used to evaluate the design for the assessment of the implementation fidelity. The design of DIVA was assessed as being 100% compliant with three out of five PDSA features (Iterative cycles, Prediction-based test of change, Small scale tests of change, Use of data over time). However, documentation was assessed to be 92% compliant and small-scale tests of change were omitted from the DIVA design. On the other hand, Table 5 also shows the gaps on each respective feature. Both tables highlight where design failure and/or implementation failure may exist, which have implications on the effectiveness of the intervention.

**Table 4:** Scores on analyses criteria

| PDSA Feature | Analyses criteria | Response score (mean) for each question |  |
| --- | --- | --- | --- |
|  |  | Design<br>(Adaptation Score) | Implementation<br>(Fidelity score) |
| Iterative cycles | Multiple cycles | 2.00 | 2.00 |
|  | Cycles linked to one another | 2.00 | 1.33 |
|  | **Composite score (%)** | **100** | **83** |
| Prediction-based test of change | Test of change | 2 | 2 |
|  | Explicit prediction articulated | 2 | 2 |
|  | **Composite score (%)** | **100** | **100** |
| Small scale tests of change | Sample size per cycle | 0 | Not applicable |
|  | Temporal duration of cycles |  |  |
|  | Number of changes tested per cycle |  |  |
|  | **Composite score (%)** | **0** | **Not applicable** |
| Use of data over time | Data collected over time | 2 | 2 |
|  | **Composite score (%)** | **100** | **100** |
| Documentation | Application of the PDSA method detailed in the reports | 2 | 1 |
|  | Documentation of each stage of the PDSA cycles | 1.67 | 1 |

| *Composite score (%)* | *92* | *50* |
| --- | --- | --- |

**Table 5:** Summary of findings and assessments

| Feature of PDSA | DIVA Design | Design gap (%) | DIVA implementation | Implementation gap (%) | Assessment |
| --- | --- | --- | --- | --- | --- |
| **Iterative cycles** | DIVA is intended to be implemented using multiple iterative cycles which are interlinked such that the 'Verify/Adjust' phases of one cycle feed into the 'Diagnose' phase of the next. | 0 | DIVA is implemented using multiple (quarterly) cycles which are linked. However, in practice, we observed that while in many cycles the 'Verify/Adjust' phases of one cycle feed into the 'Diagnose' phase of the next, there were occasions where such interlinks were interrupted. This is often due to contextual influences such as donor interests, leadership and administrative deficiencies(Eboreime, Nxumalo, *et al.*, 2018) . | 17 | Optimal theory-design consonance with mild implementation gap |
| **Prediction-based test of change** | The model tests changes in six supply and demand side determinants of health system performance. It includes tools and measures for explicitly predicting change in these determinants(NPHCDA, 2013).. | 0 | During the 'Diagnose/Intervene' phases, action plans to address health system bottlenecks are made which include quantified coverage improvement targets to be met following implementation. | 0 | Optimal theory-design-implementation consonance |
| **Small scale tests of change** | DIVA was designed to be integrated into and implemented across the existing health system. Consequently, it was not practicable to build in small tests of change as that will imply a parallel health system. | 100 | Given that the intervention was introduced and implemented at scale ab initio, the sample size remains the same in every DIVA cycle. Thus, implementation aligns with the adapted design but not with the PDSA theory. | Not applicable | Design defect, implementation fidelity not applicable |
| **Use of data over time** | The design of DIVA is to build on existing data systems. Thus, routine data from the Health Management Information System is used over time to analyse change. Given the capacity of implementers (mostly district health managers and other stakeholders from the local community), DIVA guidelines recommend use of simple descriptive statistical evaluations rather than complex analytic processes such as statistical control charts | 0 | Data was collected over time and analysed using time series charts and bargraphs. The implementation wholly complies with design. | 0 | Zero design defect with optimal implementation fidelity |
| **Documentation** | The implementation guide contains processes and tools for documenting activities, plans and lessons learned from implementation for each stage of the implementation. | 8 | There was detailed documentation of the 'Diagnose/Intervene' phases. This was less so with the 'Verify/Adjust' phases which were observed to be the weakest link in the implementation chain. | 50 | Good theory-design consonance with poor implementation fidelity |

Conversely, implementation fidelity scores were only optimal with prediction-based test of change and the use of data over time. Multiple cycles were implemented but not appropriately linked to each other. Further the application of the PDSA methods, including the four stages, were not well documented in the reports. This may imply that any observed poor performance of DIVA may have been caused by implementation failure.

## Discussion

This study interrogates the conceptual underpinnings of DIVA and its implementation fidelity in a subnational health system in Nigeria. The implementation science literature recognizes that interventions are successful only, when adapted to the local contexts (McKleroy *et al.*, 2006; Galbraith *et al.*, 2009). However, the process of modifying an intervention must not compromise its core features (theory and internal logic), which are most likely to produce the main expected outcomes. The adaptation therefore must preserve the mechanism through which the desired change in outcomes is achieved, while at the same time assuring that the interventions is a good fit to the problem being addressed (McKleroy *et al.*, 2006; Pérez *et al.*, 2015; Sundell *et al.*, 2016). Design gaps occur when the core components of the intervention are compromised during the adaptation. Implementation gaps occur when implementers fail to adhere to the execution of the design as specified by guidelines. This gap in implementation fidelity commonly results from unstable implementation environment such as evolving policies and discretionary action of implementers, among others (Eboreime, Eyles, *et al.*, 2018; Eboreime, Nxumalo, *et al.*, 2018).

Our principal finding in this study is that DIVA remains true to the principles of the PDSA approach, while excluding some components that are not germane to the objective of strengthening the planning process. One such component is the PDSA hallmark of conducting small tests of change. This is mainly because typical PDSAs are used within an organizational context such as to improve services within a clinic. DIVA, however, was designed for higher (systems) level improvement such as complex district health systems which coordinate governance and services across several communities and facilities located within defined geographical boundaries(UNICEF, 2012; Reed and Card, 2016; Eboreime, Idika, *et al.*, 2019; Eboreime, Nxumalo, *et al.*, 2019). Typically, in organizational contexts, PDSAs start small in scale and expand scale as confidence grows. However, DIVA, in Nigeria, was designed to review healthcare processes that had already been implemented and changes in each cycle were intended to address reallocation of resources or priorities rather than making changes to the processes themselves. In fact, small tests of change may disrupt existing routine systems, compromising the goal of integrated health system strengthening with implications for sustainability of the intervention (UNICEF, 2012; Eboreime *et al.*, 2017; Eboreime, Nxumalo, *et al.*, 2018). This being said, there may be potential value in applying small tests of change should DIVA be implemented in a newly designed health system or as part of holistic reforms of an existing system. That way lessons learned from these tests can inform strategies and interventions that could sustainably improve health system performance.

Another variation in DIVA compared to PDSA is the method of analysing data over time. While temporal trends are reviewed the method of analysis is descriptive, primarily using before and after bar charts. This prevents a more rigorous analysis of process variation. The adaptation was necessary because DIVA is designed to be an easy-to-use tool for decision making by health care managers/policymakers who are not trained in rules for interpreting run charts.

We also found that the implementation of DIVA varied partly from the design. Complete implementation fidelity (adherence to design) was observed in two of the five features of PDSA (prediction-based tests of change and the use of data over time). Implementation gaps were observed in two features: iterative cycles and documentation. The remaining PDSA feature, small scale tests of change, was not applicable to assess as it was omitted from the DIVA design, as discussed above. Typical PDSA cycles are iterative, with lessons learned from one cycle feeding into the ‘Plan’ phase of the cycle that follows. While this is also the design of DIVA, in practice, interruptions to these cycle linkages were observed.

In an another article (Eboreime, Nxumalo, *et al.*, 2018), using Kaplan’s Model for Understanding Success in Quality (MUSIQ) (Kaplan *et al.*, 2012), the authors described how contextual factors such as weak political will, administrative inefficiencies and donor interests partly compromised the implementation of DIVA, particularly the ‘Verify/Adjust’ phases. These deficiencies had implications on linkages between cycles on some occasions. The detailed documentation observed with the ‘Diagnose/Intervene’ phases, was not replicated with the ‘Verify/Adjust’ phases, thus reinforcing the untoward effect of these contextual factors with implications on effectiveness.

As noted in previous studies (Eboreime *et al.*, 2017; Eboreime, Nxumalo, *et al.*, 2018, 2019), DIVA is potentially effective as it was able to improve isolated performance indicators of vertical programmes such as malaria management and measles vaccination, but did not significantly improve integrated health system performance due to poor implementation of the model. This study provides further insights into possible reasons for suboptimal effectiveness of DIVA. First, a smaller 17% gap in implementing iterative cycles, then the larger 50% gap in documentation. The poor attention given to documentation could have compromised the quality of other processes, given that lessons learned from one cycle may not completely feed into subsequent cycles. This phenomenon may potentially create some form of cyclical redundancies, compromising the effectiveness of the intervention. Thus, to improve the effectiveness of DIVA in this context, more attention must be paid to documentation of processes, outcomes and lessons learned. Beyond the context in which this research was carried out, this study demonstrates that the standard principles of quality improvement interventions should inform both the design and implementation of the interventions in order to ensure effective outcomes.

The design of this study may subject the findings to some interpreter bias and subjectivity. Despite these apparent biases, we view the measures and approaches used here as practical proxies for evaluating the alignment of theory, design and implementation in the real-world.

## Conclusion

This study demonstrates how both adaptation and implementation are important for success of QI interventions (Borrelli *et al.*, 2005; Carroll *et al.*, 2007; Stirman *et al.*, 2013; El-Sadr, Philip and Justman, 2014). It also presents an approach for evaluating other QI models using Taylor’s PDSA assessment framework as a guide, which might serve to strengthen the theory behind future QI models and provide guidance on their appropriate use. This will be particularly important for improvement models like DIVA, designed for multi-contextual improvements, where different adaptations may be necessary in different countries and where implementation challenges may vary by setting.

## Data Availability

Data is available upon reasonable request

## Acknowledgements

We acknowledge the support (including permission to use data) of Nigeria’s National Primary Health Care Development Agency and the Kaduna State Primary Healthcare Development Agency, Nigeria.

